# Exploring the Potential of Large Language Models in Personalized Diabetes Treatment Strategies

**DOI:** 10.1101/2023.06.30.23292034

**Authors:** Hao Yang, Jiaxi Li, Siru Liu, Lei Du, Xiali Liu, Yong Huang, Qingke Shi, Jialin Liu

## Abstract

This study aims to explore the application of a fine-tuned model-based outpatient treatment support system for the treatment of patients with diabetes, and evaluate its effectiveness and potential value.

**Methods:** The ChatGLM model was selected as the subject of investigation and trained using the P-tuning and LoRA fine-tuning methods. Subsequently, the fine-tuned model was successfully integrated into the Hospital Information System (HIS). The system generates personalized treatment recommendations, laboratory test suggestions, and medication prompts based on patients’ basic information, chief complaints, medical history, and diagnosis data.

**Results:** Experimental testing revealed that the fine-tuned ChatGLM model is capable of generating accurate treatment recommendations based on patient information, while providing appropriate laboratory test suggestions and medication prompts. However, for patients with complex medical records, the model’s outputs may carry certain risks and cannot fully substitute outpatient physicians’ clinical judgment and decision-making abilities. The model’s input data is confined to electronic health record (EHR), limiting the ability to comprehensively reconstruct the patient’s treatment process and occasionally leading to misjudgments of the patient’s treatment goals.

**Conclusion:** This study demonstrates the potential of the fine-tuned ChatGLM model in assisting the treatment of patients with diabetes, providing reference recommendations to healthcare professionals to enhance work efficiency and quality. However, further improvements and optimizations are still required, particularly regarding medication therapy and the model’s adaptability.

## Introduction

Diabetes is a chronic metabolic disorder characterized by elevated blood glucose levels, which over time can cause serious damage to the heart, blood vessels, eyes, kidneys and nervous system. The most common type is type 2 diabetes, which accounts for about 90-95% of diabetes cases [1] and affects mainly adults. According to the World Health Organization, approximately 422 million people worldwide have diabetes, the majority in low- and middle-income countries, and 1.5 million people die each year as a direct result of diabetes [2]. The incidence and prevalence of diabetes have been steadily increasing over the last few decades [1]. As diabetes patients require long-term medication to control blood glucose levels and prevent complications [3], they can face several challenges during the treatment process, such as medication selection, dosage adjustment and management of adverse effects. Failure to address these issues in a timely manner can compromise the efficacy of medication and even pose a threat to patients’ lives [4]. Therefore, people with diabetes need timely medication advice, health education and nutrition support to help them use their medicines correctly, safely and effectively, thereby improving adherence and quality of life. To better serve patients and increase the efficiency of healthcare professionals, we aim to optimize the management of diabetes patients through the application of artificial intelligence.

With the significant success of ChatGPT in tasks related to understanding and generating human-like responses [5], large language models (LLMs) have attracted considerable attention. They have shown strong performance in various natural language processing (NLP) tasks and the ability to generalize to unfamiliar tasks, demonstrating their potential as a unified solution for natural language understanding, text generation and dialogue. Although ChatGPT has shown promising results in medical document summarization and decision support [7,8], as well as in passing the United States Medical Licensing Examination (USMLE) Step 1 and 2 [6], the exploration of these broad-domain LLMs in the medical field is still relatively limited [9]. Currently, there is a lack of specifically trained LLMs in the field of healthcare. To address this gap, we plan to fine-tune a large language model using de-identified data from diabetes patients, with the aim of exploring its application in diabetes management. In addition, harnessing the potential of large language models will open up new opportunities for medical research and practice, and drive advances and innovation in healthcare technology.

## Methods

In this study, the West China Hospital Big Data Integration Platform will be used as the data collection source [10]. We will collect EHR data from patients diagnosed with diabetes who visited the outpatient department from January 2022 to February 2022. The collected data will include information such as the patient’s department of visit, age, gender, chief complaint, present illness history, and diagnosis, which will serve as input for the model. In addition, we will obtain data on patients’ outpatient medications, laboratory tests, examinations, and physician opinions, which will be used as model outputs. Patient data with missing chief complaints and present illness history will be excluded to ensure data quality and usability. Furthermore, we will collect an additional set of data from 300 patients (visiting in March 2022) as a test set to evaluate the performance of the optimized model. Figure 1 illustrates the data collection process.

**Figure1.**
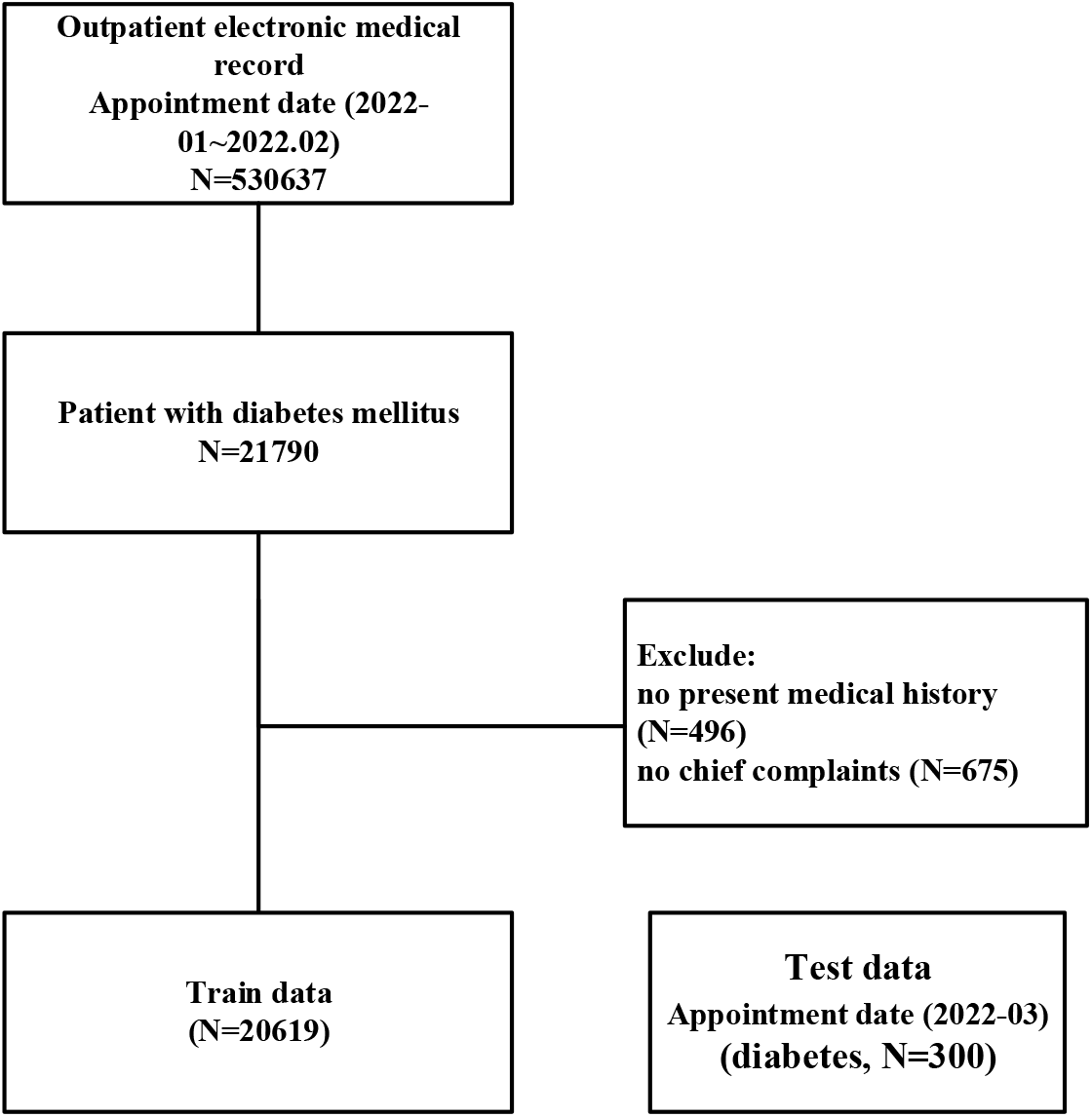
The patient selection process

### Data Pre-processing

Prior to model training, we pre-process the collected data. This includes data cleaning, standardization and anonymization. Data cleaning involves the removal of missing values, outliers and duplicates. Standardization ensures that different data elements have a consistent format and unit, which facilitates the input and output processing of the model. To protect patient privacy, we will anonymize the personal identity information of patients. For example, we will use anonymization techniques to replace or remove names and identification numbers.

### Model selection

ChatGLM is an LLM developed by Tsinghua University and Zhipu.AI [11]. ChatGLM-6B is an open-source dialogue language model based on the General Language Model (GLM) architecture, specifically designed for question answering in both Chinese and English. It consists of 6 billion parameters [11]. The open-source nature of the model is an important consideration when choosing a model. With the implementation of model quantization techniques, it can be deployed locally on consumer-grade graphics cards. The experimental environment for this study utilized a single GPU, specifically an RTX 3090 with 24GB of memory, and a CPU with 16 vCPUs (Intel(R) Xeon(R) Platinum 8350C CPU @ 2.60GHz). These hardware components were used to facilitate the training and inference processes of the ChatGLM-6B model.

### Fine-tune

We will use ChatGLM as the base model and fine-tune it for specific tasks in the field of diabetes management. For the fine-tuning process, we will employ commonly used techniques for LLMs, namely P-tuning [12] and LoRA [13]. These methods have been proven effective in adapting pre-trained language models to domain-specific tasks and improving their performance [14].

P-tuning: P-tuning is a method that automatically constructs templates for language models to perform downstream tasks [12]. It uses [unused]* tokens as continuous prompts and optimizes them with labelled data. It can achieve comparable or better performance than fine-tuning with much fewer trainable parameters. P-tuning v2 is an improved version of P-tuning that applies continuous prompts to each layer of the pre-trained model and adapts it to natural language understanding tasks [15]. In this study, we used p-tuning v2 as the technical implementation of fine-tuning (Figure 2).

**Figure 2.**
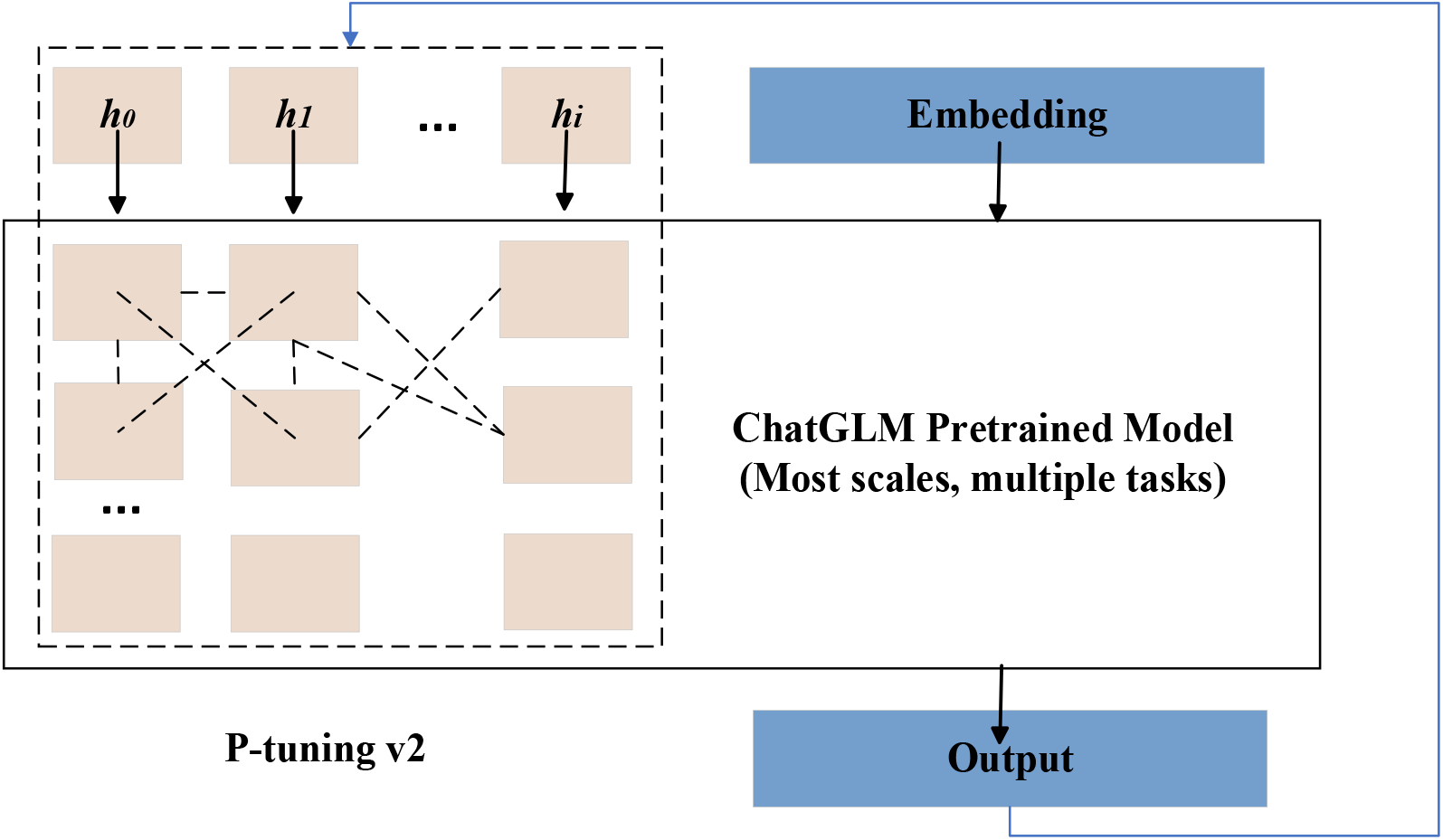
P-tuning (Each layer of the transformer requires fine-tuning of the embedding)

#### LoRA

Lora (low-rank adaptation) is a low-rank adaptation technique that allows for a significant reduction in the number of trainable parameters in downstream tasks while keeping the weights of the pre-trained model unchanged [13]. This method is similar to matrix factorization, where a trainable low-rank decomposition matrix is injected into each layer of the Transformer structure. By employing a fully connected layer, the dimensionality of the trainable layers is reduced from ‘d’ to ‘r’, and then mapped back to the original dimension ‘d’ through another fully connected layer, where ‘r’ represents the rank of the matrix, and ‘r<<d’. As a result, the computational complexity of matrix operations is reduced from ‘d×d’ to ‘d×r + r×d’, leading to a significant decrease in the number of parameters. In Lora, both parameters A and B are initialized using random Gaussian initialization (Figure 3).

**Figure3.**
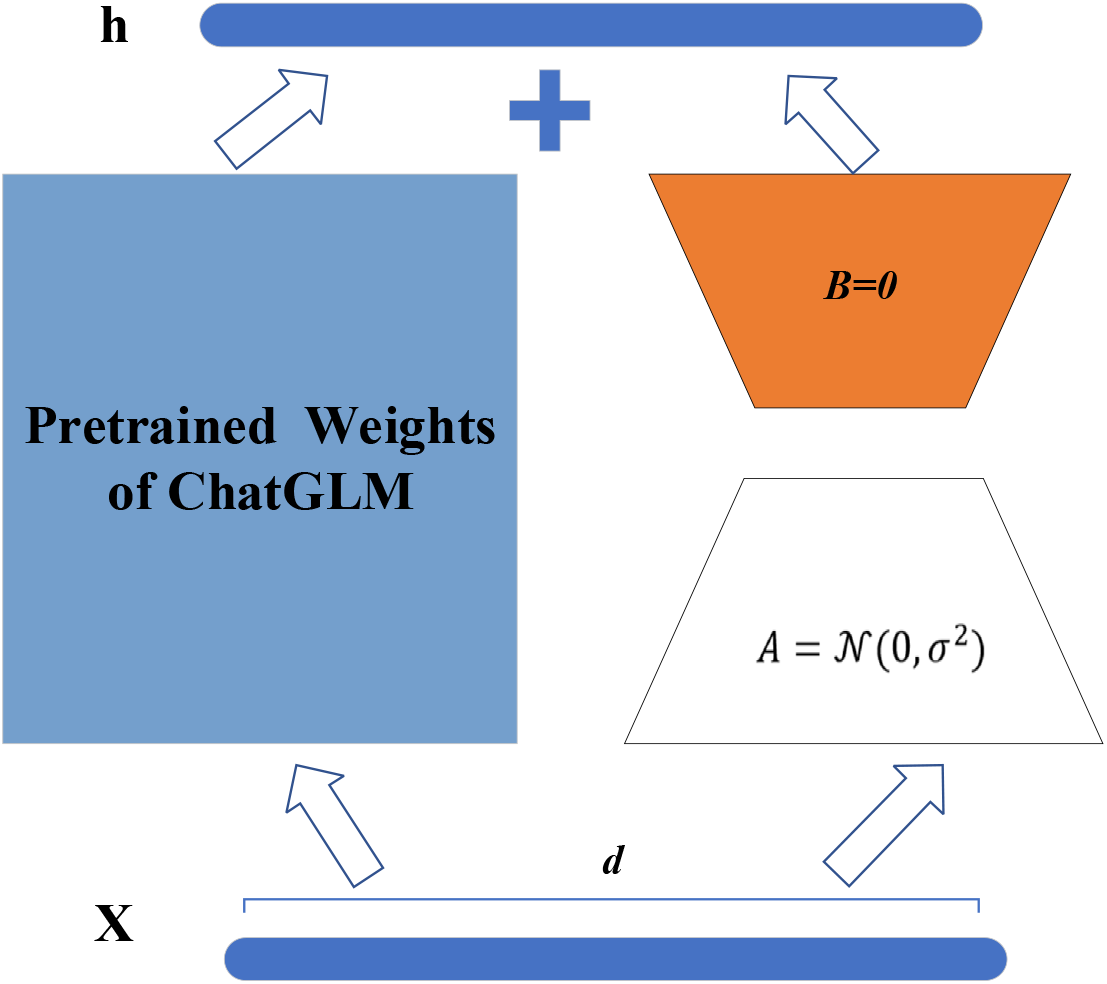
LoRA (Fine-tuning is only required for the matrices B and A)[13]

### Model metrics

#### BLEU

BLEU (Bilingual Evaluation Understudy) measures the effectiveness of a model by computing the n-gram matching between the machine-generated content and the reference standard text [16].

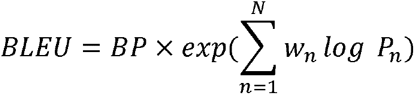

In the BLEU, the BP (Brevity Penalty) is a penalty factor that addresses the issue of shorter machine-generated content compared to the reference text. The weights, w_n_, represent the importance assigned to each n-gram matching score. P_n_ refers to the precision of the n-gram matching.

#### ROUGE

ROUGE (Recall-Oriented Understudy for Gisting Evaluation) is an evaluation metric that compares the system-generated content with manually generated reference texts. It measures the overlap between the two by counting the number of shared basic units, such as n-grams, word sequences, and word pairs, to assess the effectiveness of a model [17]. In the context of Chinese NLP tasks, ROUGE Chinese is employed for algorithm optimization. It involves transforming Chinese summaries into numeric IDs and then utilizing dynamic programming to calculate the counts of matching units, such as the longest common subsequence and N-grams. This approach allows for the computation of ROUGE-N and ROUGE-L metrics.

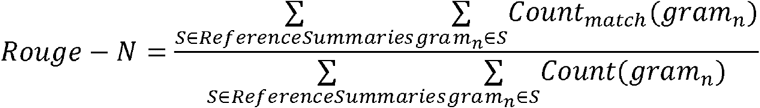

In the Roug-N, S represents the set of generated content, gram_n_ represents the n-gram, Count_match (gram_n_) indicates the number of n-grams shared between the reference and generated content, and Count (gram_n_) represents the total number of n-grams in the reference. ROUGE-N is used to measure the overlap at the n-gram level between the automatically generated text and the reference text. It is a commonly used evaluation metric.

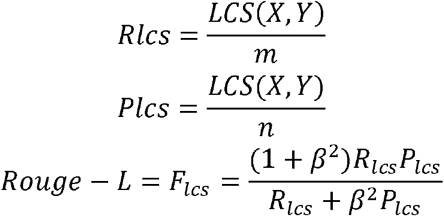

In the Rouge-L, LCS(X, Y) is the length of the longest common subsequence between X and Y. m and n are lengths of the reference summary and the automatic summary, typically measured in the number of words. R_lcs_ and P_lcs_ denote recall and precision, respectively. Finally, F_lcs_ corresponds to what we refer to as Rouge-L. In this formula, β is set to a large value, so Rouge-L primarily considers Rl_cs_.

### Physician assessment of recommendations

After the fine-tuning process, we will evaluate the model using the reserved dataset of 300 patients. The evaluation will assess the accuracy and consistency of the model in terms of diabetes treatment recommendations. We will compare the model-generated outputs and seek evaluations from three endocrinologists based on the following three categories: useful, useless, and harmful.

#### Useful

This category refers to cases where the model provides correct treatment recommendations that are helpful in clinical practice [18].

#### Useless

This category pertains to instances where the content generated by the model lacks logical coherence or specific guiding significance in certain tasks [19].

#### Harmful

This category addresses potential negative impacts and risks that the model might introduce in its application [20]. For example, it may include erroneous medication or treatment suggestions.

### Model integrated

The fine-tuned model will be integrated into the EHR system (Figure 4) to facilitate the management of diabetes patients by clinical physicians. This integration will enable the provision of personalized treatment recommendations for diabetes patients. By integrating the model with the EHR, physicians will have easy access to the system’s output, enhancing their ability to make informed decisions and develop tailored management plans for patients with diabetes.

**Figure 4.**
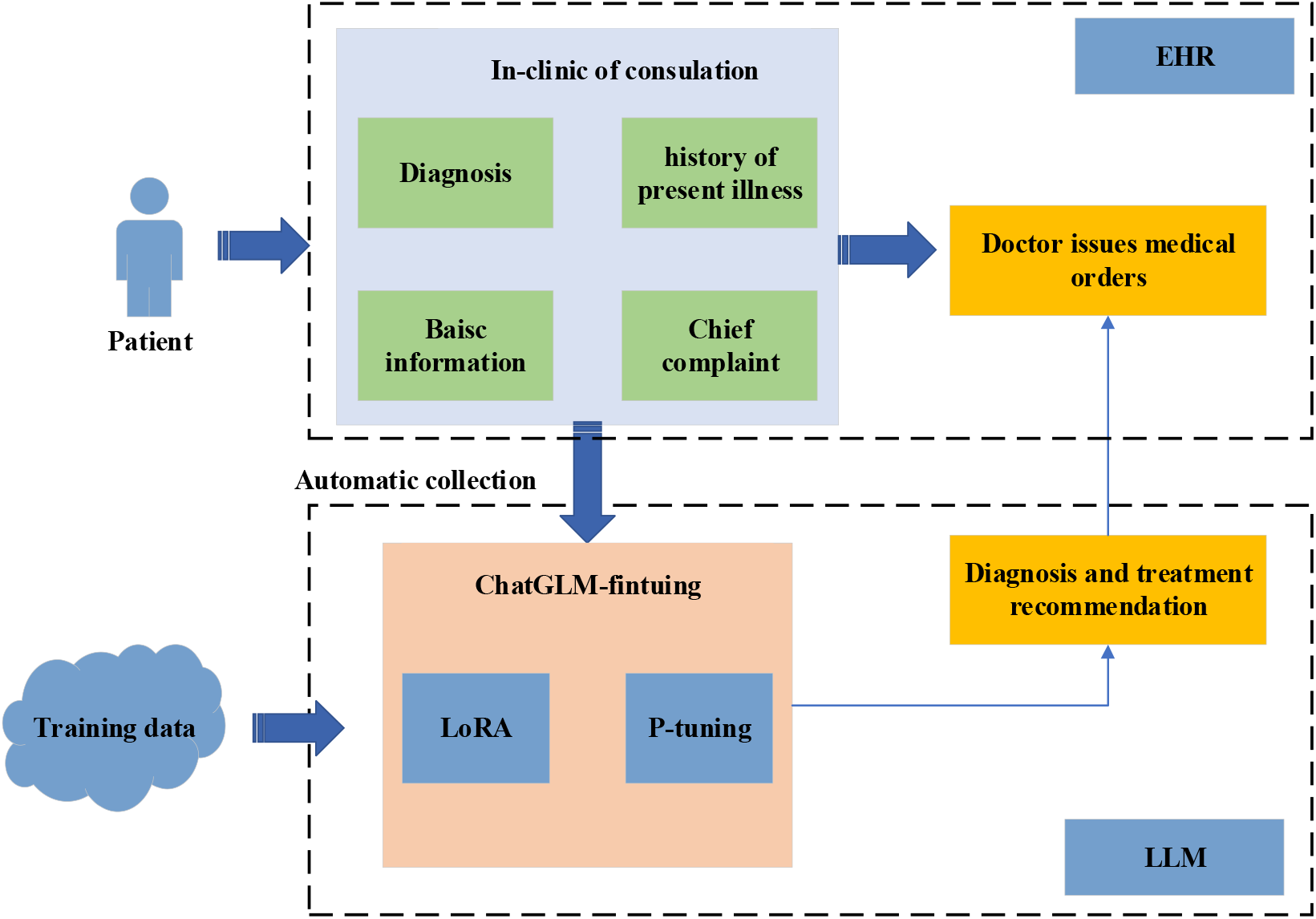
ChatGLM fine-tuning based treatment aid model for diabetes

### Ethics

Throughout the research and model development process, we will strictly adhere to ethical standards and privacy principles. Patient data will be de-identified to protect their privacy. We will ensure secure storage and transmission of the data and will only use it for model training and evaluation when necessary. In addition, this study has been approved by the Ethics Committee of West China Hospital, Sichuan University (2021-1450).

## Results

The LoRA and P-Tuning training resulted in a training loss curve for both fine-tuning methods (Figure 5). The results show that the LoRA can converge faster and achieve better fine-tuning results than P-Tuning. These results indicate that the LoRA fine-tuning method achieves significant improvements in the task of diabetes management, enhancing model performance and adaptability. It provides clinicians with more accurate and personalized guidance and recommendations.

**Figure 5.**
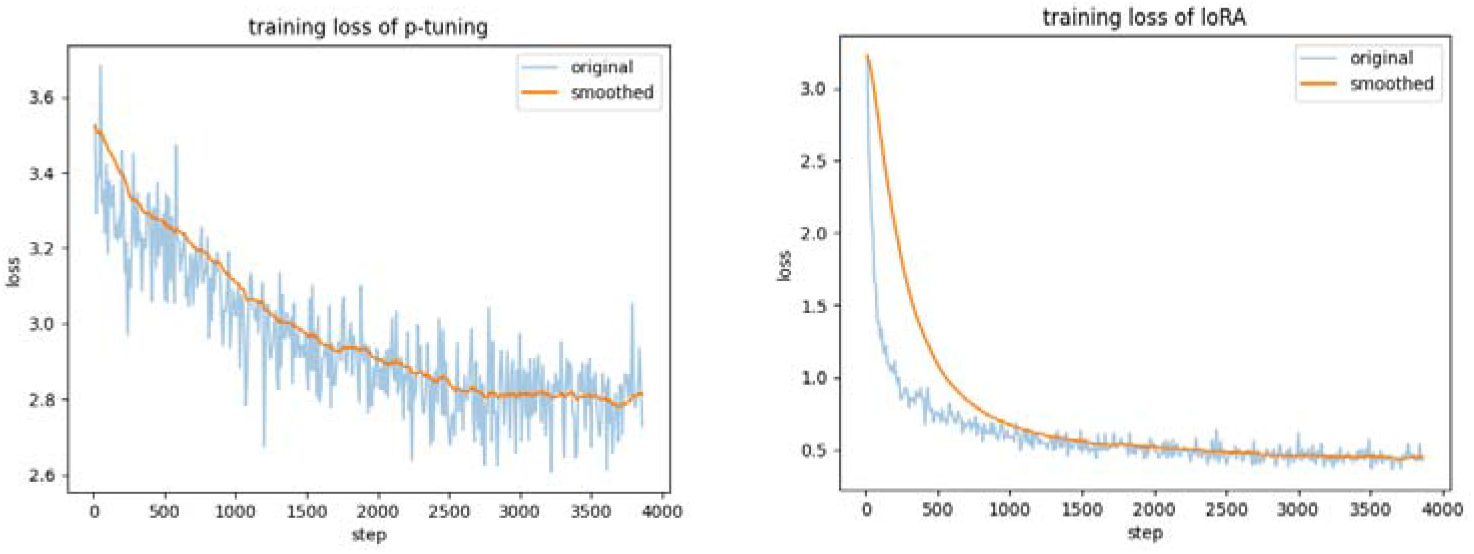
Training Loss for LoRA and P-Tuning

The results shown in Table 2, the comparison between the fine-tuned models and the original model in terms of BLEU and Rouge on the test set are as follows. The model after LoRA tuning shows a significant improvement over the original model (p < 0.05). The model after P-Tuning shows a significant improvement in the BLEU score compared to the original model (p < 0.05), while there is no improvement in the Rouge score (p > 0.05).

**Table 2.** Performance analysis of based on BLEU and Rouge scores (mean:std)

| Score | Original | P-Tuning | P (Orignal vs P-Tuning) | LoRA | P (Orignal vs LoRA) |
|---|---|---|---|---|---|
| **BLEU-4** | 42.02(0.81) | 50.22(3.47) | <0.05 | 56.54 (3.49) | <0.05 |
| **Rouge-1** | 9.40(0.04) | 9.41(0.04) | 0.7 | 33.23 (4.02) | <0.05 |
| **Rouge-2** | 0.58(0.09) | 0.49(0.08) | 0.7 | 20.37 (1.66) | <0.05 |
| **Rouge-L** | 6.18(0.07) | 6.21(0.11) | 1.0 | 28.75 (2.86) | <0.05 |

For the same question, the original model, the model after P-tuning and the LoRA show the following specific performances:

### Example

##### Input

##**Department**: Endocrinology and Metabolism Medical Unit

##**Gender**: Male

##**Age**: 65∼70 years old

##**Diagnosis**: Type 2 diabetes; hyperlipidemia

##**Chief complaint**:Diabetes follow-up visit

##**Medical history**:Diabetes for 2 more years

### Real clinical data

##### Output

**##Laboratory tests**

Biochemistry 2: Liver and kidney glucose and fat-a, glycated hemoglobin (HbA1c)

**##Medication:**

Ganjing insulin injection (3ml:300 units/pre-filled) 11 U once a day, OuTangning (5mg×7 tablets) 5 mg once a day, Atorvastatin calcium tablets (10mg×28 tablets) 10 mg once every night.

### Origin ChatGLM

##### Output

A patient with diabetes who comes for a follow-up visit needs to undergo a detailed medical history and physical examination in order to better understand the patient’s condition and develop a treatment plan. According to your description, the patient has type 2 diabetes and hyperlipidemia. Diabetes is a chronic disease that requires long-term management and treatment. Hyperlipidemia is a common metabolic disorder that can affect the patient’s cardiovascular health and increase the risk of cardiovascular disease. During the follow-up visit, the patient needs to monitor blood glucose levels and adjust them according to the control of blood glucose levels, diet, exercise, etc. In addition, the patient also needs to undergo regular blood lipid level checks and develop corresponding treatment plans based on the results of the checks.

For diabetes, common treatment options include oral medications, insulin injections, blood glucose monitoring, and dietary control. During the treatment process, patients need to actively cooperate with doctors, adhere to dietary and exercise control, and have regular follow-up visits to maintain stable and healthy blood glucose levels.

### P-tuning ChatGLM

##### Output

According to the patient’s diagnosis, he needs to be treated for both type 2 diabetes and hyperlipidemia. Here are some treatment recommendations:

1. Drug treatment: The patient needs to continue drug treatment, including oral medications and insulin. Depending on the patient’s blood glucose and blood lipid levels, the doctor may adjust the drug dosage or switch to another medication.
2. Dietary control: The patient needs to control his diet, avoid high-sugar and high-fat foods, and increase his exercise. It is recommended that patients eat high-fiber, low-fat, and low-sugar foods such as vegetables, fruits, whole grains, legumes, lean meats, fish, etc.
3. Lifestyle adjustments: The patient needs to quit smoking, limit alcohol consumption, control weight, maintain regular exercise and sleep patterns.
4. Regular follow-up visits: The patient needs to have regular follow-up visits to monitor blood glucose and blood lipid levels and adjust drug dosage or lifestyle in a timely manner.

Weight control: The patient needs to control his weight and avoid obesity to reduce the risk of hyperlipidemia.

### LoRA ChatGLM

##### Output

**##Laboratory tests**

Glycosylated hemoglobin (HbA1c)

Biochemistry 1: Liver and kidney lipase-a, Fasting insulin, 2-hour postprandial insulin, Fasting plasma glucose, 2-hour postprandial C-peptide, 2-hour postprandial plasma glucose

**##Medication:**

Gan Jing insulin injection (3ml:300 units/pre-filled) 11 U once a day, OuTangning (5mg×7 tablets) 5 mg once a day

**## Physician’s recommendation:**

Regular follow-up is recommended.

### Recommended evaluation

Table 4 shows the results of three endocrinologists’ evaluation of the 300 treatment recommendations generated by ChatGLM. The results show that more than 50% of the recommendations are clinically relevant to diabetes management, while more than 30% are of limited help to physicians. In addition, about 10% of the recommendations carried potential risks, such as incorrect treatment recommendations or overtreatment. There was no significant difference in the results of the three physician ratings (p>0.05).

**Table 4.** Physician assessment of recommendations

| Doctor | ☺useful | 😐useless | ☹harmful | P |
|---|---|---|---|---|
| #1 | 187(62.33%) | 91(30.33%) | 22(7.33%) | 0.26 |
| #2 | 172(57.33%) | 95(31.67%) | 33(11.00%) |  |
| #3 | 167(55.67%) | 108(36%) | 25(8.33%) |  |

### Integration with existing EHR

In order to seamlessly integrate ChatGLM into existing clinical workflows and EHR systems, we conducted internal testing by integrating ChatGLM into the hospital’s EHR system within an internal test environment. In this internal test environment, when a physician completes the patient’s basic information, chief complaints, history of present illness and diagnosis, the system automatically collects the information and uses a fine-tuned GLM to generate appropriate treatment recommendations. Physicians can refer to these treatment recommendations and use them as a reference when issuing medical orders and treatment suggestions to patients (Figure 6). It is important to note that these treatment recommendations serve as a reference for physicians and do not replace their decision-making process. Physicians are still required to make a comprehensive assessment based on the specific circumstances of each patient and to make final decisions based on their clinical experience and expertise.

**Figure 6.** Integration with existing EMR (Regarding the HIS system screenshots, if you require Figure 6, please contact the corresponding author.)

## Discussion

ChatGLM, developed by Tsinghua University and Zhipu.AI, is an open-source LLM for natural language understanding and generation. It is a general pre-training framework that can be effectively parameterized for different tasks. In our study, we used the ChatGLM model and trained it on data from diabetic patients.

In terms of fine-tuning, traditional fine-tuning algorithms typically require manual selection of parameters to be fine-tuned, which requires domain knowledge and expertise. In addition, LLM typically have a large number of parameters, which is an important consideration for GPU computing power and training dataset requirements. Even if only a small fraction of the parameters are frozen, forward and backward propagation computations are still required, and these computations scale linearly with the number of model parameters. As a result, large-scale models require relatively large GPU computing power and training data sets. To address these issues, our research has used the P-tuning and LoRA methods for fine-tuning training.

P-tuning is a prompt-based fine-tuning method that guides the model to produce more accurate outputs by inserting specific markers in the input. Compared to traditional fine-tuning methods, P-tuning makes better use of the knowledge of the pre-trained model and achieves better results on small datasets [21]. Moreover, P-tuning allows control over the style and content of the generated text by adjusting the prompt [22].

LoRA is a parameter-based fine-tuning method that improves performance by fine-tuning the parameters of the model [23]. Compared to traditional fine-tuning methods, LoRA makes better use of the knowledge of the pre-trained model and provides better interpretability [24]. In addition, LoRA can further improve the efficiency of a model by reducing the number of parameters through low-rank decomposition [25]. It is important to note that low-rank decomposition may introduce some loss of information as it approximates the original model parameters [13]. Therefore, when applying LoRA or other parameter reduction techniques, it is essential to strike a balance between model efficiency and performance to ensure adequate information retention and model effectiveness [26].

After repeated debugging and testing of the model, we have found that for a subset of patients with complex medical records, the model output can be potentially harmful and fail to assist healthcare professionals in their treatment. This situation can occur for the following reasons: 1) Data imbalance or sample bias: The model may have been trained on an over-representation of certain types of patient records, leading to an inadequate understanding of other types of patients. This bias can lead to inaccurate or harmful treatment recommendations for certain patients. 2) Unknown or rare scenarios: If the model encounters unfamiliar or infrequent situations during training, it may struggle to make accurate predictions or appropriate recommendations. Complex patient records often contain such unknown scenarios, rendering the model’s output ineffective. 3) Limitations of the model: The model may have inherent design or training limitations that prevent it from adequately accounting for factors specific to complex medical records. As a result, the model’s outputs may lack accuracy or reliability in these cases. 4) The size of the training data has a direct impact on the performance of large models. Therefore, it is crucial for us to explore methods to increase the scale of our dataset. [27] (supplementary material).

It is important to understand that while LLMs often perform well in many scenarios, they may have limitations when dealing with complex medical records. Therefore, model outputs should not be the sole basis for decision making. Healthcare professionals should rely on their expertise and clinical judgement, and integrate model outputs with comprehensive assessments to make informed decisions [27].

This research also has some limitations. Firstly, this work is at an early stage and contains some errors, making it unsuitable for any commercial or clinical use. The aforementioned experimental environment is limited to the experimental testing phase. Secondly, ChatGLM-6B is a GLM with only 6 billion parameters, which is significantly smaller than larger models such as ChatGPT and GPT-4, which have hundreds of billions of parameters. However, due to its smaller size, it does not require powerful computing resources and can be deployed on an intranet at a lower cost. Thirdly, the training data for this study comes from a single medical institution and is relatively small in volume. The results of the study require further validation. In future research, we will consider using larger models and incorporating more training data to improve the robustness of the results.

## Conclusion

While ChatGLM has shown promise in assisting with the treatment of diabetic patients, thorough validation, scrutiny and regulatory measures must be undertaken to ensure safety and efficacy before the model can be applied in clinical practice or commercial settings. Future research directions include developing more universally applicable medical assistance models, incorporating Reinforcement Learning from Human Feedback (RLHF) techniques for model fine-tuning, and ensuring the model’s applicability in different hospitals and real-world scenarios.

## Data Availability

All data produced in the present study are available upon reasonable request to the authors

